# Baseline α-synuclein seeding activity and disease progression in sporadic and genetic Parkinson’s disease in the PPMI cohort

**DOI:** 10.1101/2024.09.27.24311107

**Authors:** Jackson G. Schumacher, Xinyuan Zhang, Eric A. Macklin, Jian Wang, Armin Bayati, Johannes M. Dijkstra, Hirohisa Watanabe, Michael A. Schwarzschild, Marianna Cortese, Xuehong Zhang, Xiqun Chen

**Author notes:** Denotes equal contribution (Jackson G. Schumacher, Xinyuan Zhang). Denotes senior author (Marianna Cortese, Xuehong Zhang, Xiqun Chen). Correspondence to: Xiqun Chen, Department of Neurology, Massachusetts General Hospital and Harvard Medical School, Boston, MA 02129, USA.

## Abstract

**Background:** α-Synuclein (α-syn) seed amplification assays (SAAs) have shown remarkable potential in diagnosing Parkinson’s disease (PD). Using data from the Parkinson’s Progression Markers Initiative (PPMI) cohort, we aimed to test whether baseline α-syn seeding activity was associated with disease progression in sporadic PD, *LRRK2*-associated PD (*LRRK2* PD), and *GBA*-associated PD (*GBA* PD).

**Methods:** We analyzed 7 years of motor, non-motor, and cognitive assessments and 5 years of dopamine transporter imaging along with baseline α-syn SAA results from 564 PPMI participants (n=332 sporadic PD, 162 *LRRK2* PD, and 70 *GBA* PD) using linear mixed-effects models, adjusted for potential confounders, to test whether baseline α-syn SAA positivity (n=315 sporadic PD, 111 *LRRK2* PD, and 66 *GBA* PD) was associated with PD progression.

**Results:** While non-statistically significant, there was a trend towards faster motor decline in participants with α-syn SAA positive *LRRK2* PD compared to those with α-syn SAA negative *LRRK2* PD (MDS-UPDRS III points per year: 2.39 (95% confidence interval: 1.86 – 2.92) vs. 1.76 (0.93 – 2.60); difference=0.63 (−0.29 – 1.55, p=0.18). There was no difference in motor decline between α-syn SAA positive and α-syn SAA negative participants with sporadic PD (2.46 (2.20 – 2.72) vs. 2.39 (1.36 – 3.42); difference=0.07 (−0.99 – 1.12), p=0.90) or *GBA* PD (2.67 (1.91 – 3.44) vs. 2.40 (−0.18 – 4.99); difference=0.27 (−2.42 – 2.96), p=0.84). No statistically significant differences were seen in the progression of non-motor symptoms, cognition, or DAT imaging.

**Conclusions:** We found no statistically significant associations between baseline α-syn seeding activity and PD progression among manifest patients in the PPMI cohort. Future studies are needed to further investigate relationships among baseline α-syn seeding activity, disease heterogeneity, disease stage, and PD progression.

## Introduction

Parkinson’s disease (PD) is a progressive neurodegenerative disorder characterized by motor symptoms such as bradykinesia, resting tremors, stiffness, and impaired balance and coordination, along with non-motor symptoms, including cognitive decline, mood changes, and autonomic dysfunction.^1^ Pathologically, PD is primarily characterized by abnormally aggregated α-synuclein (α-syn) and degeneration of dopaminergic neurons in the substantia nigra (SN). A substantial body of evidence indicates that the aggregation of monomeric α-syn into pathogenic oligomeric confirmations and fibrils contributes to dopaminergic neurodegeneration and broader pathological processes in PD.^2^ Once these pathogenic aggregates form, they have been shown to propagate between cells, amplifying and accelerating disease pathology. The propagation of pathogenic α-syn aggregates is currently the prevailing theory behind the progressive nature of PD.^2–6^

Techniques to detect pathogenic α-syn seeds, collectively known as α-syn seed amplification assays (SAAs), leverage the ability of α-syn seeds to induce aggregation of recombinant α-synuclein monomers to enable their detection even at extremely low concentrations.^7^ α-Syn SAAs have shown high sensitivity and specificity when differentiating between sporadic PD and healthy controls.^7^ Additionally, multiple cross-sectional studies have shown that α-syn seeding activity is associated with more severe motor dysfunction, cognitive impairment, brain atrophy, and reduced connectivity in PD, as well as worse clinical outcomes in Alzheimer’s disease (AD).^8–11^ As such, α-syn SAAs have had a remarkable impact on the field and have prompted calls for a biological definition and staging system of PD.^12,13^ However, whether α-syn SAAs may provide information on PD progression remains unknown.

While the majority of PD cases are sporadic, variants in leucine-rich repeat kinase 2 (*LRRK2*) and glucocerebrosidase (*GBA*) are among the most common genetic risk factors for the disease.^14,15^ Previous studies have shown that pathogenic *LRRK2* gain-of-function variants are associated with slower PD progression, while *GBA* loss-of-function variants are associated with faster PD progression.^16,17^ Interestingly, a study by the Parkinson’s Progression Markers Initiative (PPMI reported significantly lower rates of α-syn SAA positivity in carriers of the *LRRK2* G2019S variant than those with sporadic PD.^18^ Here, we analyzed 7 years of clinical assessments and 5 years of imaging data from 564 PPMI participants with baseline α-syn SAA results to determine whether α-syn seeding activity was associated with progression in sporadic, *LRRK2*, and *GBA* PD.

## Methods

### Analysis design and participants

This cohort analysis is based on data collected by the PPMI study and includes participants from the Parkinson’s Disease Cohort and Genetic Registry. We excluded participants who met enrollment criteria but had scans without evidence of dopamine deficiency (SWEDD)(n=66). Genome-wide sequencing data from PPMI was used to categorize these participants into three groups: participants with sporadic PD (non-carriers of any known pathogenic mutations), participants with *LRRK2* PD (carriers of *LRRK2* G2019S, R1441C/G+M1646T, and N2081D/N14D variants), and participants with *GBA* PD (carriers of *GBA* E326K, R502C, A495P, and N409S variants). We excluded participants with *SNCA* (n=38), *PRKN* (n=9), *PINK1* (n=1), *PARK7* (n=1), and *VPS35* (n=1) mutations as well as those with both *LRRK2* and *GBA* mutations (n=24).

The data analyzed was collected by PPMI between the study’s inception, July 7, 2010, and May 24, 2024. More detailed information about PPMI’s mission and inclusion criteria, as well as sources and methods of participant selection, can be found on the PPMI website (www.ppmi-info.org/access-data-specimens/download-data). The data was last accessed using the PPMI portal on June 24, 2024.

### Clinical and SAA data

Participants underwent clinical assessments and imaging by PPMI-associated clinicians at baseline and during subsequent follow-up visits as previously described.^19,20^ Baseline refers to when participants were recruited into the cohort. Although PPMI has up to 14 years of Movement Disorder Society Unified Parkinson’s Disease Rating Scale Part I and III (MDS-UPDRS I and III) and Montreal Cognitive Assessment (MoCA) data, the data beyond 7 years is sparse, particularly for participants with *LRRK2* and *GBA* PD. As such, we analyzed the first 7 years (mean follow-up = 5.81 years) of MDS-UPDRS I and III and MoCA data, as well as 5 years (mean follow-up = 3.98 years) of dopamine transporter (DAT) imaging with single-photon emission computed tomography (DAT-SPECT) as measures of PD-related non-motor decline, motor decline, cognitive decline, and loss of DAT, respectively.^21–23^ Only “off” medication state MDS-UPDRS III assessments were included. The number of observations collected vs. time for MDS-UPDRS I and III, MoCA, and DAT-SPECT data is shown in Supplemental Figure 1.

Cerebrospinal fluid (CSF) was collected from participants (n=564) at baseline and analyzed by the PPMI team using the 24-hour (n=61) and 150-hour (n=505) Amprion α-syn SAAs.^18,24^ While there is some interprotocol variability, we used data from both the 24-hour and 150-hour Amprion α-syn SAAs as they have comparable protocols with the exception of timeframe. Methods for the 24-hour Amprion α-syn SAA can be found in the PPMI database.

### Statistics

All participants with genome-wide sequencing data, a PD diagnosis, and baseline α-syn SAA results at the time of data access were included in our analysis. Our statistical analysis plan was developed *a priori* and refined *post hoc*. All statistical analyses were performed using SAS® software version 9.4, and all figures were created using R software version 4.4.1.

A linear mixed-effects model was used to estimate the yearly rate of change in MDS-UPDRS I and III score, MoCA score, and DAT-SPECT specific binding ratio (SBR) for α-syn SAA positive and negative participants. Given previously reported differences in progression between participants with sporadic, *LRRK2*, and *GBA* PD, as well as reported differences in α-syn SAA positivity, we estimated separate rates of progression for participants with α-syn SAA positive and negative sporadic, *LRRK2*, and *GBA* PD.^14–16^ Each outcome was modeled independently.

Models assumed participant-level random intercepts and slopes with unstructured covariance and heteroscedastic variance between groups. All models were adjusted for baseline demographic characteristics (age, sex, race [two levels], ethnicity [two levels], and education [three levels] (all self-reported by participants)), time since original diagnosis at baseline, and baseline score, as well as their interaction with time as a fixed effect. We also adjusted for the levodopa equivalent daily dosage (LEDD) at the time of each visit. Estimates were obtained for an average participant at an LEDD of zero. A significance level of α=0.05 was used for all analyses. Confounding variables were identified based on a review of existing literature and modeled according to a directed acyclic graph (DAG)(Supplemental Figure 2). Exposure and adjustment variables were treated as fixed effects to account for their consistent influence across all participants. All model residuals were assessed by Q-Q plots for normality assumption, residuals vs. fitted plots for heteroscedastic assumption, and residuals vs. predictor plots for linearity assumption. The variance inflation factor (VIF) was used to check for multicollinearity, and variables with VIF >10 would result in revising the DAG and statistical model.

## Results

We analyzed longitudinal data from 564 PPMI participants, including 332 with sporadic PD, 162 with *LRRK2* PD, and 70 with *GBA* PD. Baseline demographic and clinical characteristics for each group categorized by α-syn SAA result are shown in Table 1. Interestingly, across sporadic, *LRRK2*, and *GBA* PD, α-syn SAA negative participants were older at disease onset (Table 1).

**Table 1.** Baseline demographic and clinical characteristics for sporadic, *LRRK2*, and *GBA* PD categorized by α-synuclein seed amplification assay result. Data is shown as n (%) or mean (standard deviation). Statistical analysis comparing group characteristics was not performed. Abbreviations: PD, Parkinson’s disease; LRRK2, leucine-rich repeat kinase 2; GBA, glucocerebrosidase; MDS-UPDRS, Movement Disorder Society Unified Parkinson’s Disease Rating Scale; MoCA, Montreal Cognitive Assessment; DAT-SPECT, dopamine transporter imaging with single-photon emission computed tomography; SBR, specific binding ratio.

|  | Sporadic PD |  | LRRK2 PD |  | GBA PD |  |
| --- | --- | --- | --- | --- | --- | --- |
|  | Positive SAA | Negative SAA | Positive SAA | Negative SAA | Positive SAA | Negative SAA |
| N | 315 | 17 | 111 | 51 | 66 | 4 |
| Age at baseline (years) | 61.8 (9.8) | 67.2 (11.0) | 59.6 (9.0) | 68.1 (6.9) | 61.4 (9.0) | 66.3 (10.3) |
| Years since original diagnosis | 0.68 (1.04) | 0.83 (0.77) | 2.43 (2.04) | 2.98 (2.13) | 2.24 (2.18) | 0.98 (0.62) |
| Male sex | 208 (66.0) | 11 (64.7) | 63 (56.8) | 20 (39.2) | 40 (60.6) | 3 (75.0) |
| Race |  |  |  |  |  |  |
| White | 283 (89.8) | 17 (100) | 100 (90.1) | 43 (84.3) | 63 (95.5) | 4 (100) |
| Asian (Including Native Hawaiian or other Pacific Islander) | 8 (2.5) | 0 | 0 | 0 | 0 | 0 |
| Black | 7 (2.2) | 0 | 0 | 0 | 0 | 0 |
| Multiracial | 10 (3.2) | 0 | 7 (6.3) | 8 (15.7) | 2 (3.0) | 0 |
| Other | 5 (1.6) | 0 | 3 (2.7) | 0 | 1 (1.5) | 0 |
| Not Reported | 2 (0.6) | 0 | 1 (0.9) | 0 | 0 | 0 |
| Hispanic or Latino ethnicity | 7 (2.2) | 0 | 19 (17.1) | 16 (31.4) | 2 (3.0) | 0 |
| Education |  |  |  |  |  |  |
| Less than 12 years | 20 (6.3) | 0 | 10 (9.0) | 15 (29.4) | 4 (6.1) | 0 |
| 12-16 years | 191 (60.6) | 9 (52.9) | 50 (45.0) | 21 (41.2) | 30 (45.5) | 2 (50.0) |
| Greater than 16 years | 104 (33.0) | 8 (47.1) | 51 (45.9) | 15 (29.4) | 32 (48.5) | 2 (50.0) |
| Relatives with Parkinson's disease |  |  |  |  |  |  |
| Parent | 37 (11.7) | 0 | 34 (30.6) | 23 (45.1) | 12 (18.2) | 1 (25.0) |
| Other | 55 (17.5) | 0 | 37 (33.3) | 25 (49.0) | 13 (19.7) | 0 |
| MDS-UPDRS III score at baseline | 20.1 (8.9) | 19.1 (5.3) | 20.1 (10.1) | 17.5 (7.9) | 25.2 (11.3) | 20.0 (13.0) |
| MDS-UPDRS I score at baseline | 4.25 (3.06) | 6.29 (5.17) | 5.62 (4.05) | 6.12 (5.02) | 5.68 (3.90) | 2.75 (2.75) |
| MoCA score at baseline | 27.2 (2.3) | 26.8 (2.1) | 26.6 (2.8) | 25.1 (3.1) | 26.4 (2.7) | 26.2 (2.6) |
| DAT-SPECT SBR at baseline |  |  |  |  |  |  |
| Caudate | 1.98 (0.52) | 1.86 (0.69) | 1.92 (0.56) | 1.89 (0.46) | 1.85 (0.69) | 1.59 (0.82) |
| Putamen | 0.81 (0.26) | 0.95 (0.54) | 0.74 (0.22) | 0.90 (0.35) | 0.73 (0.29) | 1.02 (0.95) |

Participants with α-syn SAA positive sporadic PD (n=315; MDS-UPDRS III points per year: 2.46 (95% confidence interval: 2.20 – 2.72)) progressed similarly to those with α-syn SAA negative sporadic PD (n=17; 2.39 (1.36 – 3.42); difference=0.07 (−0.99 – 1.12), p=0.90)(Figure 1 and Table 2). While the difference was not statistically significant, there was a trend toward faster motor decline in participants with α-syn SAA positive *LRRK2* PD (n=111; 2.39 (1.86 – 2.92)) compared to those with α-syn SAA negative *LRRK2* PD (n=51; 1.76 (0.93 – 2.60); difference=0.63 (−0.29 – 1.55), p=0.18). There was no statistically significant difference between participants with α-syn SAA positive *GBA* PD (n=66; 2.67 (1.91 to 3.44)) and those with α-syn SAA negative *GBA* PD (n=4; 2.40 (−0.18 to 4.99); difference=0.27 (−2.42 – 2.96), p=0.84).

**Figure 1.**
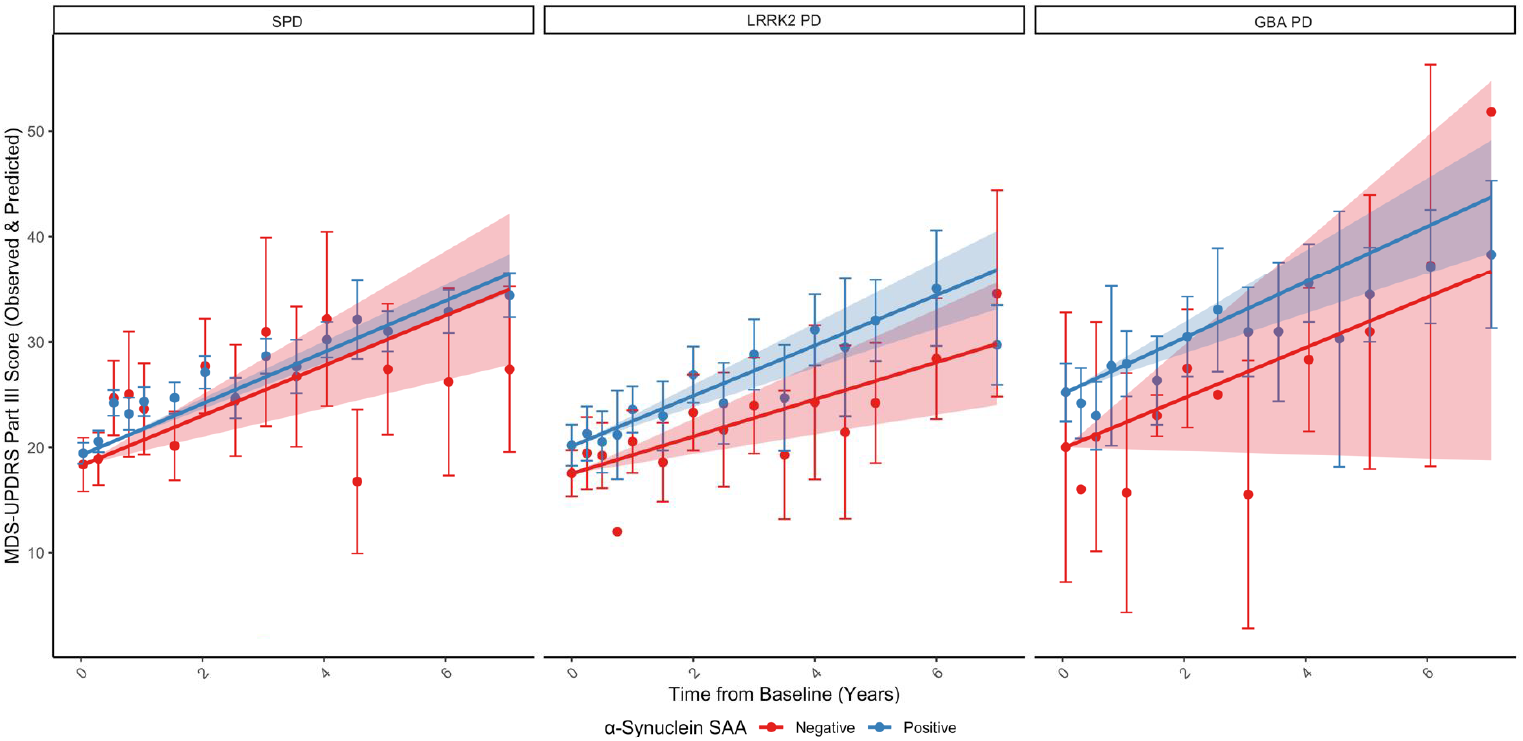
Change in MDS-UPDRS III for participants with sporadic, *LRRK2*, and *GBA* PD categorized by α-synuclein seed amplification assay result. Observed and predicted change in MDS-UPDRS III adjusted for age, time since diagnosis, sex, race, ethnicity, years of education, levodopa equivalent daily dosage, and baseline score in individuals with α-synuclein seed amplification assay positive and negative sporadic (n=315 vs. 17), *LRRK2* (n=111 vs. 51), and *GBA* PD (n=66 vs. 4). Comparing the difference in change to those in the α-syn SAA negative group as reference. Data points represent the mean observed value at the corresponding time point. Lines represent the predicted value over time after adjustment. Error bars and shading represent 95% confidence intervals for observed and predicted values. Abbreviations: PD, Parkinson’s disease; LRRK2, leucine-rich repeat kinase 2; GBA, glucocerebrosidase; α-synuclein SAA, α-synuclein seed amplification assay; MDS-UPDRS III, Movement Disorder Society Unified Parkinson’s Disease Rating Scale Part III

**Table 2.**
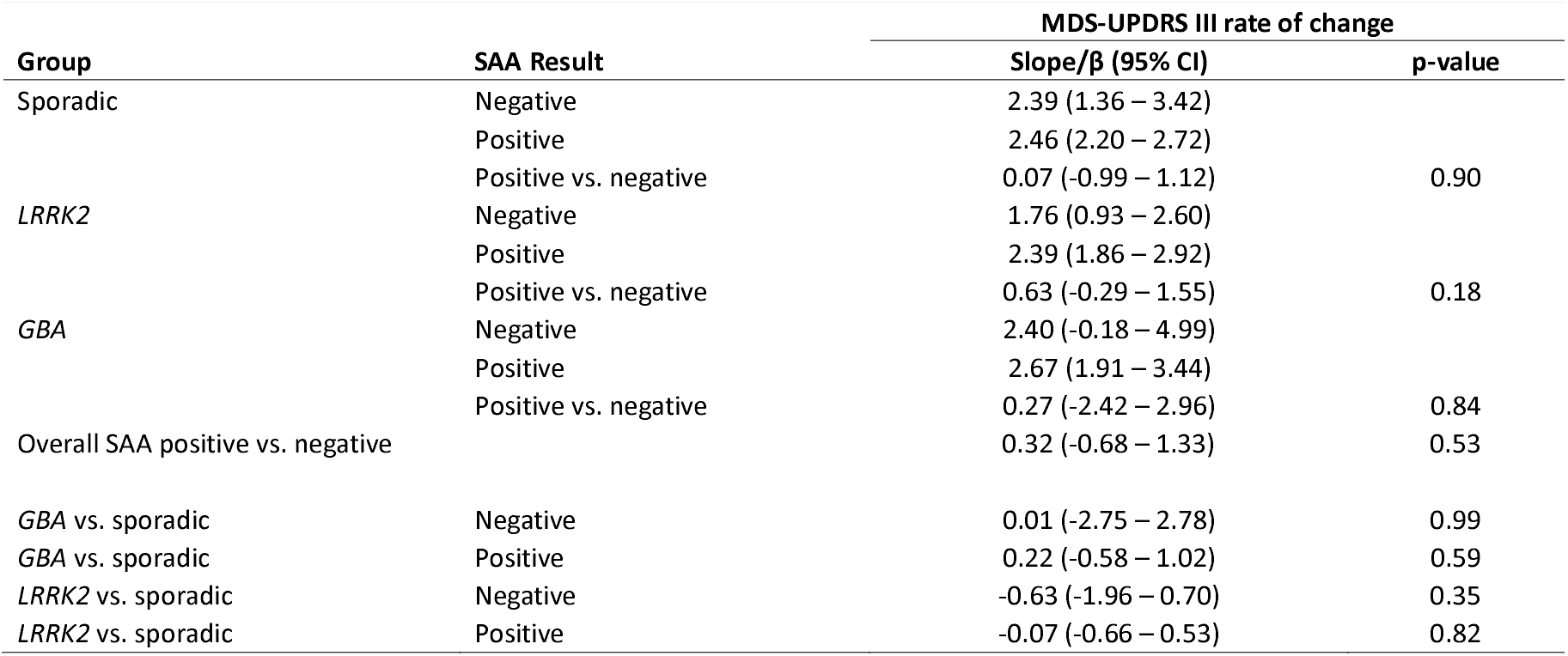
Change in MDS-UPDRS III for participants with sporadic, *LRRK2*, and *GBA* PD categorized by α-synuclein seed amplification assay result. Slopes (unit per year) were estimated from linear mixed models. Fixed effects included time * (genetic form * SAA results, baseline MDS-UPDRS III, baseline age, years since original diagnosis to baseline, sex, race [two levels], ethnicity [two levels], and education [three levels]) and LEDD. Participant-level random intercepts and slopes with unstructured covariance, heteroscedastic by genetic form. Abbreviations: PD, Parkinson’s disease; LRRK2, leucine-rich repeat kinase 2; GBA, glucocerebrosidase; α-synuclein SAA, α-synuclein seed amplification assay; MDS-UPDRS III, Movement Disorder Society Unified Parkinson’s Disease Rating Scale Part III

There was no difference in the rate of motor decline between participants with α-syn SAA positive *LRRK2* PD and their α-syn SAA positive sporadic PD counterparts (difference=−0.07 (−0.66 – 0.53); p=0.82). While there was no statistically significant difference, there was a trend towards slower motor decline in α-syn SAA negative *LRRK2* PD compared to those with α-syn SAA negative sporadic PD (difference=−0.63 (−1.96 – 0.70); p=0.35). Similarly, there was no statistically significant difference between α-syn SAA positive or negative *GBA* PD and their sporadic PD counterparts (p>0.05).

Additionally, we obtained similar results in our comparison of the rate of change in MDS-UPDRS I scores (Supplemental Figure 3 and Supplemental Table 1). The rate of change in MoCA scores was limited in all participants (Supplemental Figure 4 and Supplemental Table 1). We also found no statistically significant difference in our comparison of the rate of DAT-SPECT SBR loss (Supplemental Figures 5 and 6 and Supplemental Table 1).

The median time to dopaminergic medication initiation for participants with sporadic PD is shown in Supplemental Table 2. There were not enough participants with *LRRK2*, or *GBA* PD enrolled prior to initiation of dopaminergic medication to analyze the time to initiation.

## Discussion

The development of α-syn SAAs has transformed our ability to detect α-syn seeds and diagnose PD.^7,18^ Making use of MDS-UPDRS I and III, MoCA, DAT-SPECT, and baseline α-syn SAA data from the PPMI cohort, we found that there was no statistically significant association between baseline α-syn seeding activity and disease progression in sporadic, *LRRK2*, and *GBA* PD. However, consistent across clinical measures, α-syn SAA positive participants exhibited a numerically higher estimated rate of progression compared to their α-syn SAA negative counterparts. In particular, there was a non-statistically significant trend toward faster motor decline in α-syn SAA positive LRRK2 PD compared to α-syn SAA negative LRRK2 PD (p=0.18).

In the sporadic PD group, the overwhelming majority of participants were α-syn SAA positive (315/332 [95%]), potentially reflecting the central role of α-syn aggregation in PD pathophysiology. While prior studies have suggested that α-syn SAA negativity may indicate an atypical or misdiagnosed form of PD, our analysis did not show significant differences in progression rates.^12,13^ Given the limited number of α-syn SAA-negative participants, this finding should be interpreted with caution. Notably, the upper bound of the confidence interval indicates that participants with α-syn SAA positive sporadic PD may progress up to 1 point per year faster than their α-syn SAA negative counterparts. Nevertheless, based on a minimal clinically important difference of 4.63 points, our results are still sufficiently precise to exclude any clinically meaningful difference over at least 4 years.^27^ Future studies with larger sample sizes and additional biomarkers, such as tau or amyloid pathology, may help clarify whether α-syn SAA negative sporadic PD represents a distinct PD subtype or are a result of technical limitations of the assay.

Similarly, we found no significant differences in progression rates for *GBA* PD between α-syn SAA positive and negative participants. Given the few α-syn SAA negative *GBA* PD participants in our samples (4/70 [6%]), our estimates were not sufficiently precise to exclude a potentially clinically meaningful difference over 2 years. However, the estimated difference was quite modest. Given that *GBA* mutations are strongly associated with α-syn pathology, α-syn SAA positivity may be a less distinguishing factor in this population.^18,28^

Participants with α-syn SAA positive *LRRK2* PD exhibited a numerically faster rate of motor decline than their α-syn SAA negative counterparts; however, this difference did not reach statistical significance. The upper bound of the confidence interval indicates that participants with α-syn SAA positive LRRK2 PD may progress up to 1.5 points per year faster than their α-syn SAA negative counterparts, excluding any clinically meaningful difference over at least 3 years. A similar trend toward slower motor decline was observed in α-syn SAA negative *LRRK2* PD compared to α-syn SAA negative sporadic PD. In addition, there was a trend towards or statistically significant slower cognitive decline in *LRRK2* PD compared to sporadic PD in α-syn SAA positive and negative cases, respectively. These findings in general align with previous studies indicating that *LRRK2* PD may progress slower.^16,18,29^ Notably, the proportion of α-syn SAA positive individuals in *LRRK2* PD was substantially lower than in sporadic PD (111/162 [69%] vs. 315/332 [95%]). Future investigations should address whether the lower rates of α-syn SAA positivity and the slower disease progression observed in *LRRK2* PD are related.

We leverage rigorously assembled longitudinal and α-syn SAA data from the PPMI cohort along with a statistical model that stringently accounts for variability in disease progression both within and between participants. Overall, our findings suggest that α-syn SAA positivity is not a robust predictor of PD progression across clinical and imaging measures in this cohort. However, the absence of significant findings does not imply a lack of biological relevance but rather reflects the true nature of our data within the limitations of this study. Despite the overall high quality of the PPMI dataset, there was substantial person-to-person variability in progression rates that current prognostics cannot predict. While some meaningful differences could be excluded, our estimates were limited by imbalanced sample sizes between α-syn SAA positive and α-syn SAA negative groups. Further, compared to the overall long course of PD, a 7-year follow-up period may not be long enough to accurately address PD progression. While MoCA is complementary to MDS-UPDRS, not all individuals with PD will have cognitive deficits, and if these deficits do manifest, they typically do so later in the disease. Moreover, α-syn pathology in brain tissues may be influenced by factors beyond what is detectable in CSF collected through lumbar puncture. Additionally, while we adjusted for key confounding variables, there may still be other confounders, e.g., co-existing medical conditions and lifestyle factors, that may contribute to PD progression. Lastly, the cohort primarily contains participants of European descent, and, as such, our results may not be generalizable to non-European populations.

Our study provides fresh insight into the relationship between α-syn SAAs and PD progression. While we found no statistically significant association between α-syn SAA positivity and the rate of PD progression, the relationships among baseline α-syn seeding activity, disease heterogeneity, disease stage, and PD progression warrant further investigation. Importantly, participants with α-syn SAA negative PD still exhibit symptomatic progression, and the presence of α-syn seeding activity at baseline may not fully capture the dynamic nature of disease progression and neurodegeneration. As such, future studies should investigate longitudinal changes of α-syn seeding activity together with complementary biomarkers for PD progression, including among pre-manifest individuals.

## Data Availability

Data used in the preparation of this article is openly available to qualified researchers, and in the case of whole genome sequencing data, it is available upon request. Data was obtained in June 2024 from the Parkinson’s Progression Markers Initiative (PPMI) database (www.ppmi-info.org/access-data-specimens/download-data, RRID:SCR00_6431). For up-to-date information on the study, visit www.ppmi-info.org. All other resources and codes used in this study can be found at https://doi.org/10.5281/zenodo.14984600.

https://www.ppmi-info.org/access-dataspecimens/download-data

https://www.ppmi-info.org

https://doi.org/10.5281/zenodo.14984600

## Contributors

Conceptualization: JGS, XYZ, EAM, JW, AB, JMD, HW, MAS, MC, XHZ, and XC; Data Curation: JGS and XYZ; Methodology, Software, and Formal Analysis: JGS, XYZ, EAM, JW, JMD, HW, MAS, MC, XHZ, and XC; Visualization: JGS, and XYZ; Data Validation: JGS, XYZ, EAM, JW, AB, MC, and XC; Writing – Original Draft: JGS, XYZ, and XC; Writing – Review & Editing: JGS, XYZ, EAM, JW, AB, JMD, HW, MAS, MC, XHZ, and XC; Supervision: EAM, MAS, MC, XHZ, and XC; Funding Acquisition: XC. JS, XYZ, EAM, JW, AB, MC and XC have all had direct access to and have verified the underlying data. All authors have read and approved the final version of the manuscript.

## Declaration of interests

JGS, XYZ, JW, AB, JMD, MAS, MC, XHZ, and XC declare no conflicts of interest.

EAM received payments to his institution from AI Therapeutics, Biohaven, Calico Therapeutics, Denali Therapeutics, ITB-Med, Janssen, Lilly, Mitsubishi Tanabe Pharma America, Neurizon, Prilenia Therapeutics, Revalesio, Seelos Therapeutics, UCB/Ra Pharma, Woolsey. He received compensation for serving on scientific advisory boards for Annexon, Bial Biotech, Chase Therapeutics, HillHurst, Merck; for serving on steering committees for Biogen, UCB; and for serving on data monitoring committees for Argenx, NeuroSense, Novartis, Sanofi.

HW received payments from Takeda Pharmaceuticals, AbbVie, Kyowa Kirin, Sumitomo Pharma, Eisai, and Ono Corporation for work unrelated to this study.

## Acknowledgements

This research was funded in part by Aligning Science Across Parkinson’s No. ASAP-237603 through the Michael J. Fox Foundation for Parkinson’s Research (MJFF) and by the National Institute of Health through the National Institute of Neurological Disorders and Stroke grants R01NS102735 and 5R01NS126260. The authors would like to thank PPMI – a public-private partnership – funded by the Michael J. Fox Foundation for Parkinson’s Research and funding partners, including 4D Pharma, Abbvie, AcureX, Allergan, Amathus Therapeutics, Aligning Science Across Parkinson’s, AskBio, Avid Radiopharmaceuticals, BIAL, BioArctic, Biogen, Biohaven, BioLegend, BlueRock Therapeutics, Bristol-Myers Squibb, Calico Labs, Capsida Biotherapeutics, Celgene, Cerevel Therapeutics, Coave Therapeutics, DaCapo Brainscience, Denali, Edmond J. Safra Foundation, Eli Lilly, Gain Therapeutics, GE HealthCare, Genentech, GSK, Golub Capital, Handl Therapeutics, Insitro, Jazz Pharmaceuticals, Johnson & Johnson Innovative Medicine, Lundbeck, Merck, Meso Scale Discovery, Mission Therapeutics, Neurocrine Biosciences, Neuron23, Neuropore, Pfizer, Piramal, Prevail Therapeutics, Roche, Sanofi, Servier, Sun Pharma Advanced Research Company, Takeda, Teva, UCB, Vanqua Bio, Verily, Voyager Therapeutics, the Weston Family Foundation and Yumanity Therapeutics.

