## Supplemental Tables 1 and 2 for "Baseline α-synuclein seeding activity and disease progression in sporadic and genetic Parkinson’s disease in the PPMI cohort"

|  |  | **MDS-UPDRS I** |  | **MoCA** |  | **Caudate SBR** |  | **Putamen SBR** |  |
| --- | --- | --- | --- | --- | --- | --- | --- | --- | --- |
| **Group** | **SAA** | **Slope (95% CI)** | **p-value** | **Slope (95% CI)** | **p-value** | **Slope (95% CI)** | **p-value** | **Slope (95% CI)** | **p-value** |
| Sporadic | Negative | 0.54 (0.21, 0.87) |  | -0.34 (-0.66, -0.01) |  | -0.12 (-0.17, -0.07) |  | -0.05 (-0.08, -0.03) |  |
|  | Positive | 0.54 (0.46, 0.61) |  | -0.18 (-0.25, -0.11) |  | -0.13 (-0.14, -0.11) |  | -0.06 (-0.07, -0.06) |  |
|  | Positive vs. negative | 0 (-0.34, 0.33) | 0.99 | 0.16 (-0.17, 0.48) | 0.35 | -0.01 (-0.06, 0.04) | 0.78 | -0.01 (-0.04, 0.01) | 0.33 |
| *LRRK2* | Negative | 0.27 (0.07, 0.48) |  | 0.03 (-0.14, 0.19) |  | -0.11 (-0.14, -0.08) |  | -0.06 (-0.08, -0.05) |  |
|  | Positive | 0.36 (0.23, 0.49) |  | -0.05 (-0.16, 0.05) |  | -0.13 (-0.15, -0.11) |  | -0.07 (-0.08, -0.06) |  |
|  | Positive vs. negative | 0.08 (-0.15, 0.31) | 0.48 | -0.08 (-0.27, 0.10) | 0.37 | -0.02 (-0.06, 0.02) | 0.27 | -0.01 (-0.03, 0.01) | 0.57 |
| *GBA* | Negative | 0.19 (-0.34, 0.72) |  | -0.19 (-0.71, 0.32) |  | -0.11 (-0.23, 0.01) |  | -0.06 (-0.10, -0.02) |  |
|  | Positive | 0.51 (0.36, 0.66) |  | -0.28 (-0.43, -0.13) |  | -0.14 (-0.18, -0.11) |  | -0.07 (-0.08, -0.06) |  |
|  | Positive vs. negative | 0.32 (-0.23, 0.87) | 0.25 | -0.09 (-0.62, 0.45) | 0.75 | -0.03 (-0.16, 0.09) | 0.62 | -0.01 (-0.05, 0.03) | 0.66 |
| Overall SAA positive vs. negative | | 0.13 (-0.09, 0.36) | 0.25 | 0 (-0.22, 0.22) | 0.99 | -0.02 (-0.07, 0.03) | 0.39 | -0.01 (-0.03, 0.01) | 0.30 |
| *GBA* vs. sporadic | Negative | -0.35 (-0.97, 0.27) | 0.26 | 0.14 (-0.46, 0.74) | 0.64 | 0.01 (-0.12, 0.14) | 0.90 | -0.01 (-0.05, 0.04) | 0.76 |
| *GBA* vs. sporadic | Positive | -0.03 (-0.19, 0.14) | 0.74 | -0.10 (-0.26, 0.06) | 0.22 | -0.02 (-0.05, 0.02) | 0.36 | 0 (-0.02, 0.01) | 0.53 |
| *LRRK2* vs. sporadic | Negative | -0.27 (-0.65, 0.12) | 0.18 | 0.36 (0.01, 0.72) | 0.047 | 0.01 (-0.05, 0.07) | 0.77 | -0.01 (-0.04, 0.02) | 0.45 |
| *LRRK2* vs. sporadic | Positive | -0.18 (-0.33, -0.03) | 0.022 | 0.13 (0, 0.25) | 0.05 | -0.01 (-0.03, 0.02) | 0.70 | -0.01 (-0.02, 0.01) | 0.47 |

**Supplemental Table 1: Change in MDS-UPDRS I, MoCA, and DAT-SPECT SBR for participants with sporadic, LRRK2, and GBA PD categorized by α-synuclein seed amplification assay result**

Slopes (unit per year) were estimated from linear mixed models. Fixed effects included time * (genetic form * SAA results, baseline measure for each corresponding marker, baseline age, years since original diagnosis to baseline, sex, race [two levels], ethnicity [two levels], and education [three levels]) and LEDD. Participant-level random intercepts and slopes with unstructured covariance, heteroscedastic by genetic form.

Abbreviations: PD, Parkinson's disease; LRRK2, leucine-rich repeat kinase 2; GBA, glucocerebrosidase; MDS-UPDRS, Movement Disorder Society Unified Parkinson's Disease Rating Scale; MoCA, Montreal Cognitive Assessment; DAT-SPECT, dopamine transporter imaging with single-photon emission computed tomography; SBR, specific binding ratio.

|  | **Sporadic PD** |
| --- | --- |
| α-syn SAA- | 1.583 (2.25) |
| α-syn SAA+ | 1.583 (1.53) |

**Supplemental Table 2: Median time to dopaminergic medication initiation**

Data is shown as the median (95% CI or standard error from a Kaplan-Meier estimate) time to dopaminergic medication initiation in years.

Abbreviations: PD, Parkinson's disease; α-syn SAA, α-synuclein seed amplification assay
