## Supplemental Figures 1 to 6 for "Baseline α-synuclein seeding activity and disease progression in sporadic and genetic Parkinson’s disease in the PPMI cohort"

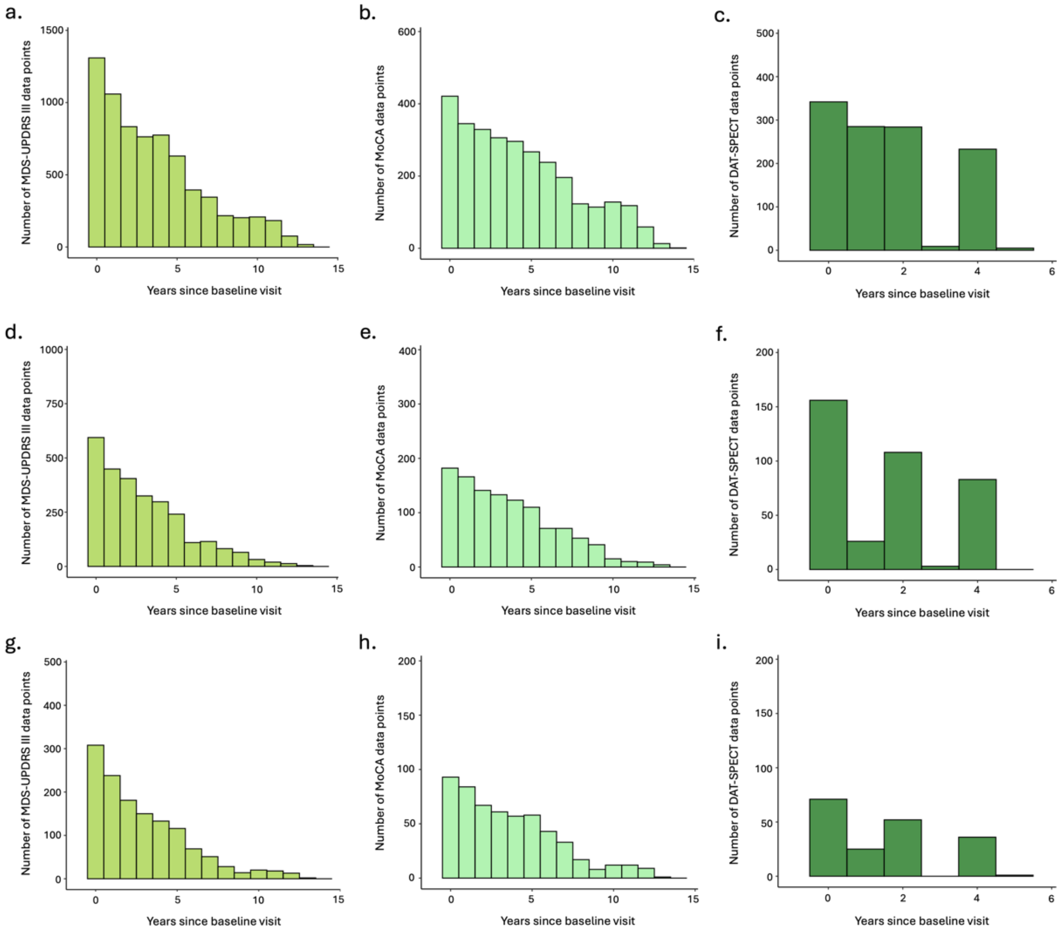


**Supplemental Figure 1. Distribution of MDS-UPDRS I and III, MoCA, and DAT-SPECT data for sporadic, LRRK2, and GBA PD**

The number of MDS-UPDRS I and III data points at each time point for sporadic PD (**a**), LRRK2 PD (**d**), and GBA PD (**g**). The number of MoCA data points at each time point (0–14 years) for sporadic PD (**b**), LRRK2 PD (**e**), and GBA PD (**h**). The number of DAT-SPECT data points available at each time point for sporadic PD (**c**), LRRK2 PD (**f**), and GBA PD (**i**).

Abbreviations: PD, Parkinson's disease; LRRK2, leucine-rich repeat kinase 2; GBA, glucocerebrosidase; MDS-UPDRS, Movement Disorder Society Unified Parkinson's Disease Rating Scale; MoCA, Montreal Cognitive Assessment; DAT-SPECT, dopamine transporter imaging with single-photon emission computed tomography; SBR, specific binding ratio.

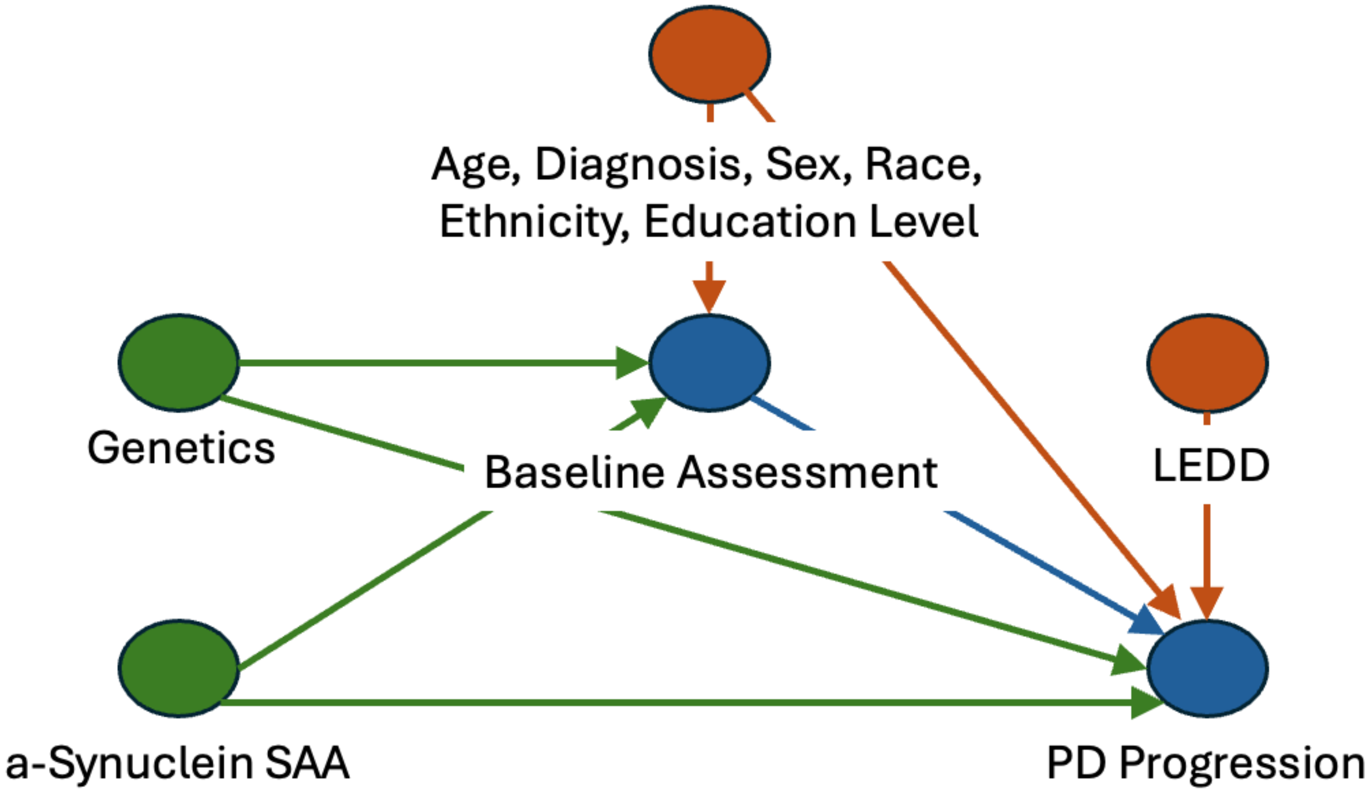


**Supplemental Figure 2. Directed acyclic graph for the identification of confounding variables**
Abbreviations: α-Synuclein SAA, α-synuclein seed amplification assay; LEDD, levodopa equivalent daily dosage; PD, Parkinson's disease

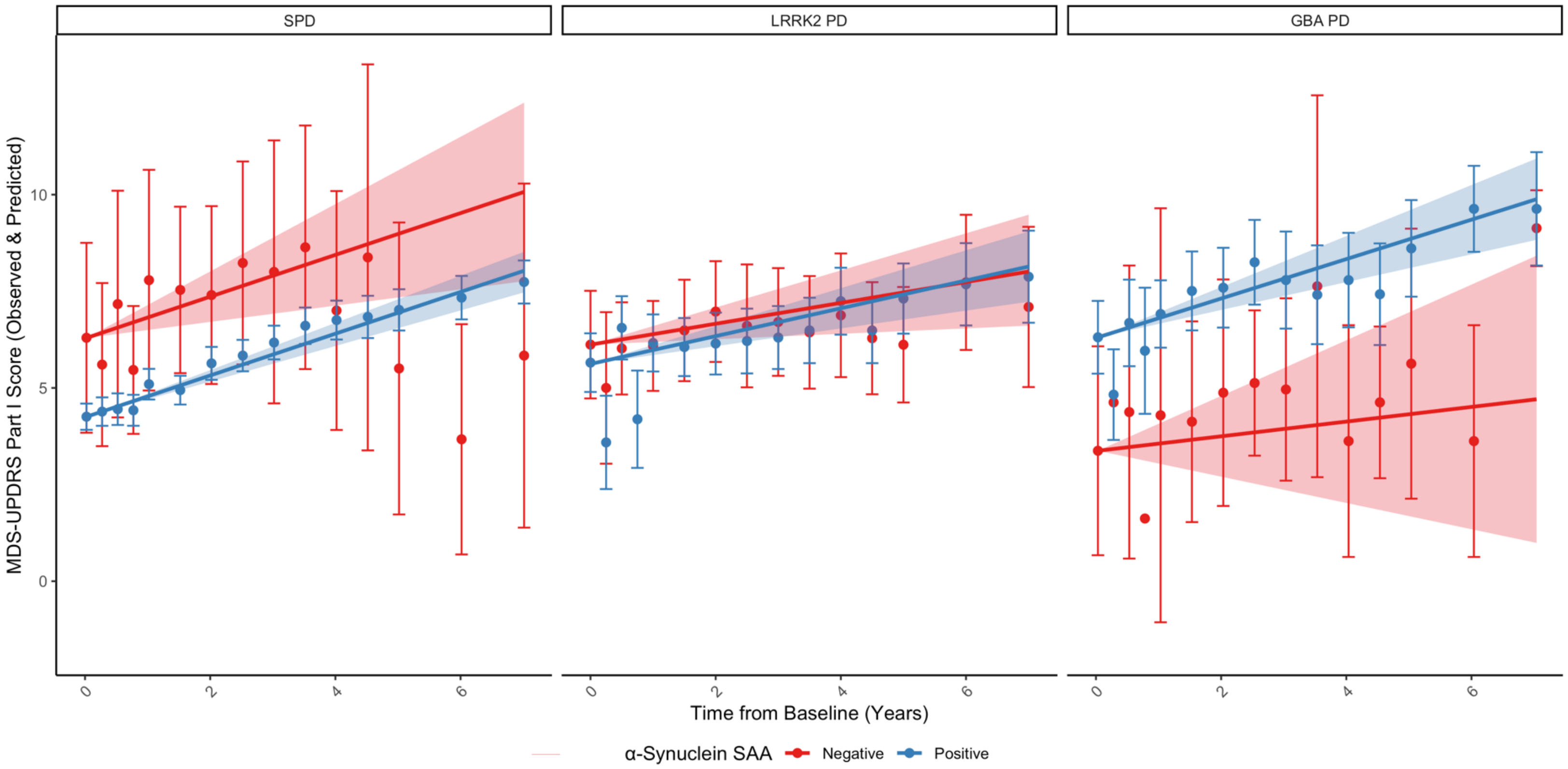


**Supplemental Figure 3. Change in MDS-UPDRS I for participants with sporadic, *LRRK2*, and *GBA* PD categorized by α-synuclein seed amplification assay result**

Observed and predicted change in MDS-UPDRS I adjusted for age, time since diagnosis, sex, race, ethnicity, years of education, levodopa equivalent daily dosage, and baseline score in individuals with sporadic, *LRRK2*, and *GBA* PD categorized by α-synuclein seed amplification assay result. Comparing the difference in change to those in the α-syn SAA negative group as reference. Data points represent the mean observed value at the corresponding time point. Lines represent the predicted value over time after adjustment. Error bars and shading represent 95% confidence intervals for observed and predicted values.

Abbreviations: PD, Parkinson's disease; LRRK2, leucine-rich repeat kinase 2; GBA, glucocerebrosidase; α-synuclein SAA, α-synuclein seed amplification assay; MDS-UPDRS I, Movement Disorder Society Unified Parkinson's Disease Rating Scale Part I

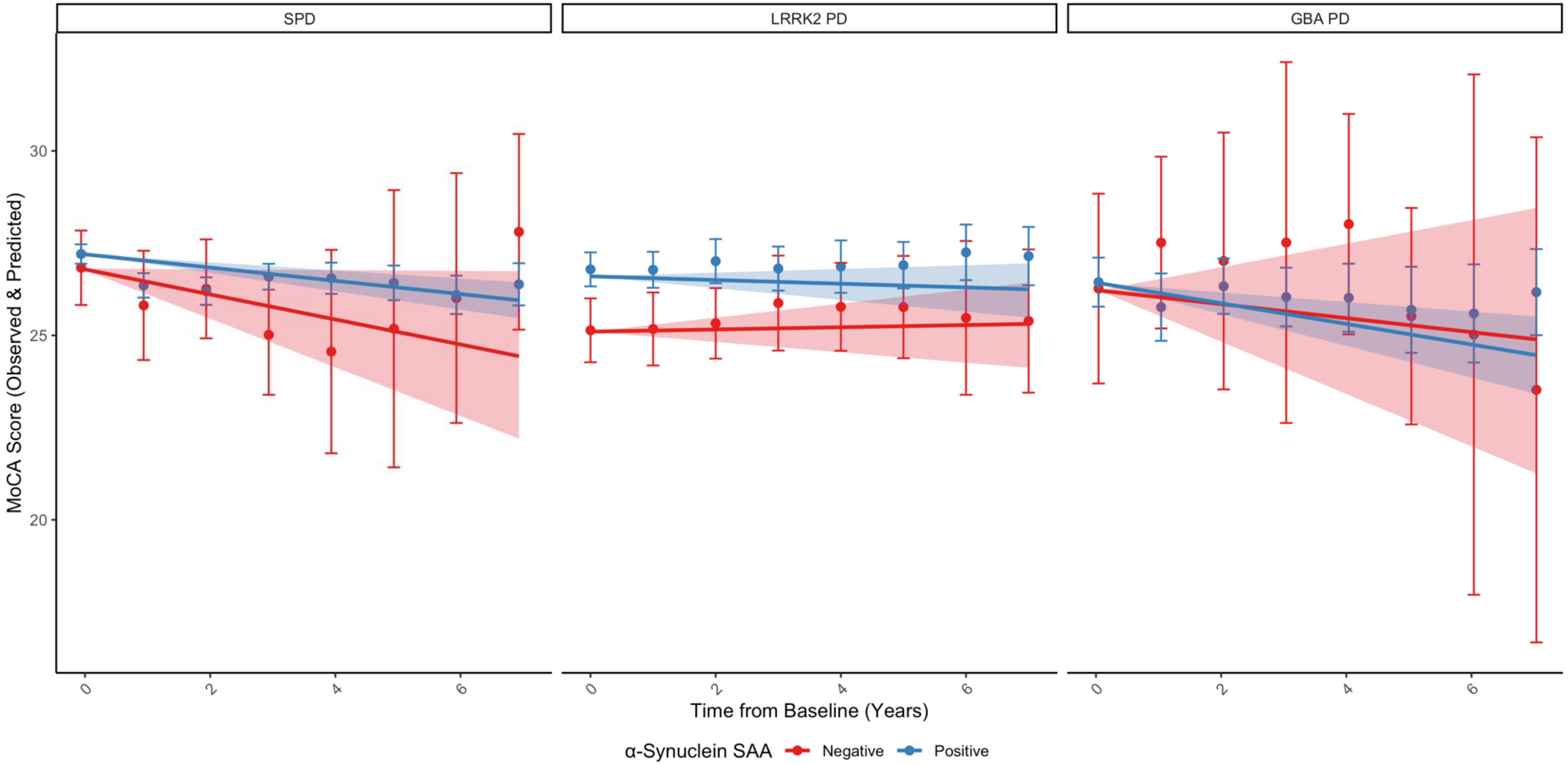


**Supplemental Figure 4. Change in MoCA for participants with sporadic, *LRRK2*, and *GBA* PD categorized by α-synuclein seed amplification assay result**

Observed and predicted change in MoCA adjusted for age, time since diagnosis, sex, race, ethnicity, years of education, levodopa equivalent daily dosage, and baseline score in individuals with sporadic, *LRRK2*, and *GBA* PD categorized by α-synuclein seed amplification assay result. Comparing the difference in change to those in the α-syn SAA negative group as reference. Data points represent the mean observed value at the corresponding time point. Lines represent the predicted value over time after adjustment. Error bars and shading represent 95% confidence intervals for observed and predicted values.

Abbreviations: PD, Parkinson's disease; LRRK2, leucine-rich repeat kinase 2; GBA, glucocerebrosidase; α-synuclein SAA, α-synuclein seed amplification assay; MoCA, Montreal Cognitive Assessment

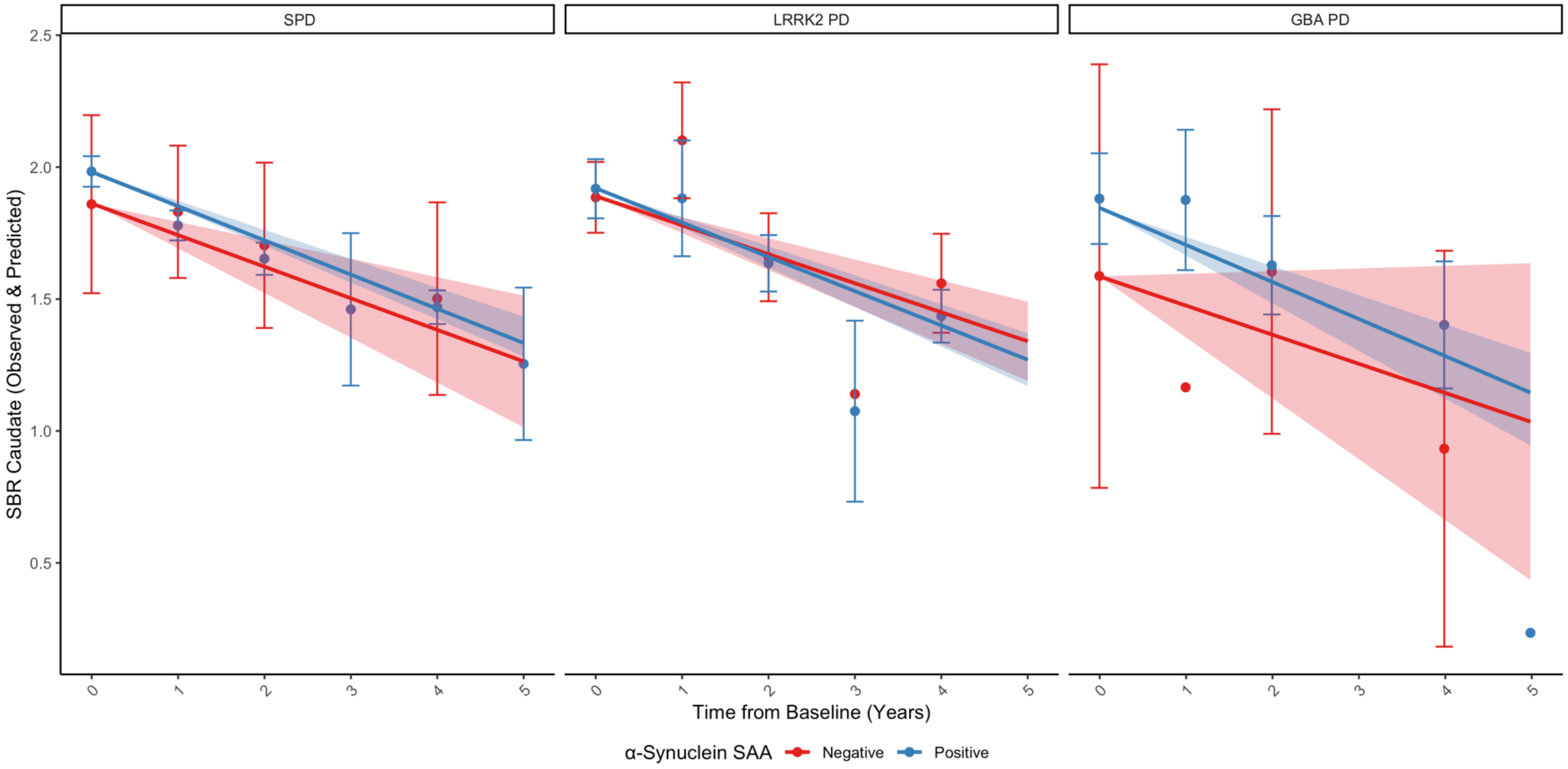


**Supplemental Figure 5. Change in DAT-SPECT caudate SBR for participants with sporadic, *LRRK2*, and *GBA* PD categorized by α-synuclein seed amplification assay result**

Observed and predicted change in DAT-SPECT caudate SBR adjusted for age, time since diagnosis, sex, race, ethnicity, years of education, levodopa equivalent daily dosage, and baseline score in individuals with sporadic, *LRRK2*, and *GBA* PD categorized by α-synuclein seed amplification assay result. Comparing the difference in change to those in the α-syn SAA negative group as reference. Data points represent the mean observed value at the corresponding time point. Lines represent the predicted value over time after adjustment. Error bars and shading represent 95% confidence intervals for observed and predicted values.

Abbreviations: PD, Parkinson's disease; LRRK2, leucine-rich repeat kinase 2; GBA, glucocerebrosidase; α-synuclein SAA, α-synuclein seed amplification assay; DAT-SPECT, dopamine transporter imaging with single-photon emission computed tomography; SBR, specific binding ratio

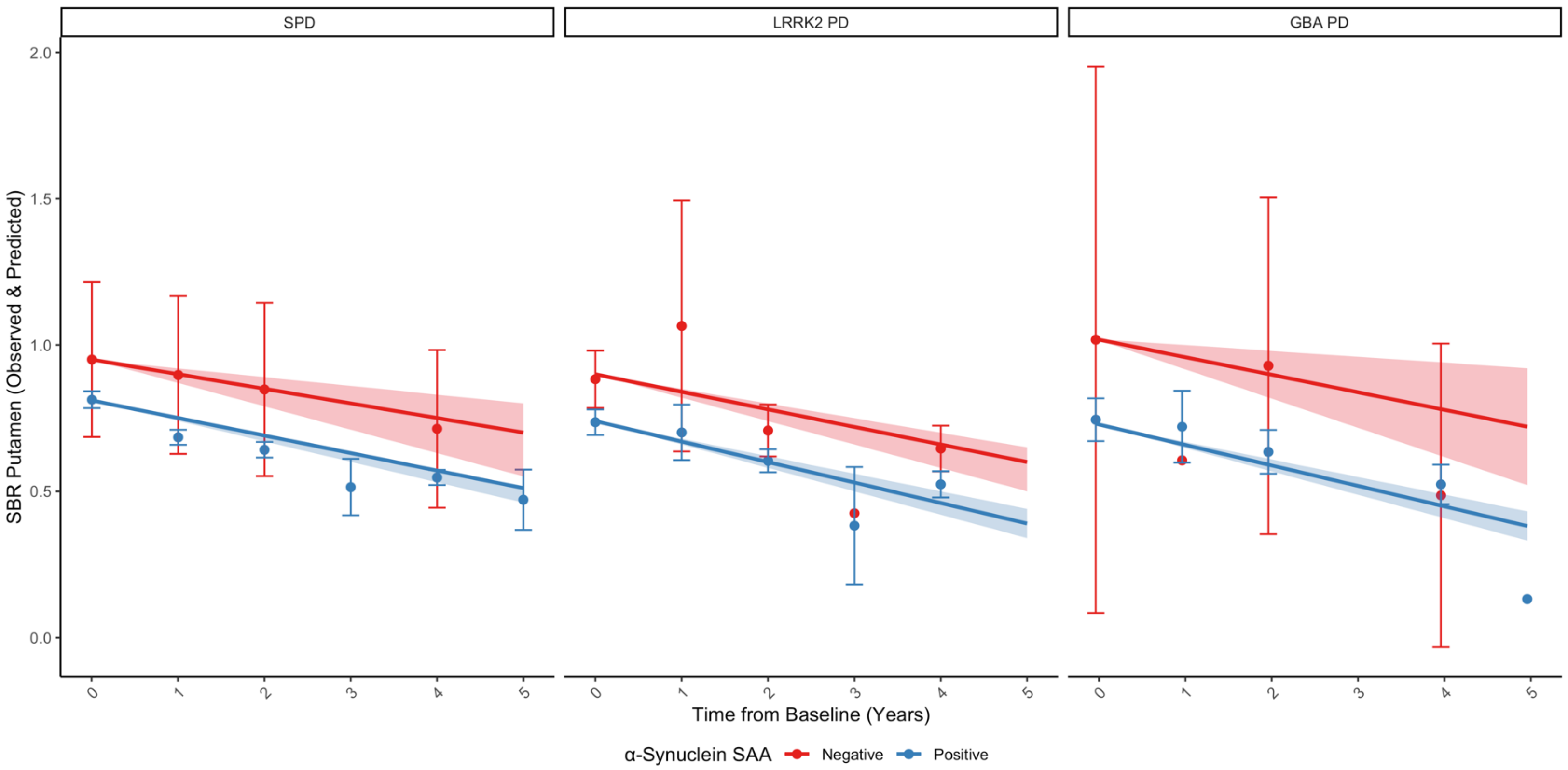


**Supplemental Figure 6. Change in DAT-SPECT putamen SBR for participants with sporadic, *LRRK2*, and *GBA* PD categorized by α-synuclein seed amplification assay result**

Observed and predicted change in DAT-SPECT putamen SBR adjusted for age, time since diagnosis, sex, race, ethnicity, years of education, levodopa equivalent daily dosage, and baseline score in individuals with sporadic, *LRRK2*, and *GBA* PD categorized by α-synuclein seed amplification assay result. Comparing the difference in change to those in the α-syn SAA negative group as reference. Data points represent the mean observed value at the corresponding time point. Lines represent the predicted value over time after adjustment. Error bars and shading represent 95% confidence intervals for observed and predicted values.

Abbreviations: PD, Parkinson's disease; LRRK2, leucine-rich repeat kinase 2; GBA, glucocerebrosidase; α-synuclein SAA, α-synuclein seed amplification assay; DAT-SPECT, dopamine transporter imaging with single-photon emission computed tomography; SBR, specific binding ratio
